# Learning the Mental Health Impact of COVID-19 in the United States with Explainable Artificial Intelligence

**DOI:** 10.1101/2020.07.19.20157164

**Authors:** Indra Prakash Jha, Raghav Awasthi, Ajit Kumar, Vibhor Kumar, Tavpritesh Sethi

**Affiliations:** Indraprastha Institute of Information Technology, Delhi (IIIT-D); XLRI - Xavier School of Management - Jamshedpur

**Keywords:** Mental Health, Covid-19, Bayesian Network, Machine Learning

## Abstract

**Importance:** COVID-19 pandemic has deeply affected the health, economic, and social fabric of nations. Identification of individual-level susceptibility factors may help people in identifying and managing their emotional, psychological, and social well-being.

**Objective:** This work is focused on learning a ranked list of factors that could indicate a predisposition to a mental disorder during the COVID pandemic.

**Data Sources and Study design:** In this study, We have used a survey of 17764 adults in the USA at different age groups, genders, and socioeconomic statuses.

**Methods:** Through initial statistical analysis followed by Bayesian Network inference, we have identified key factors affecting Mental health during the COVID pandemic. Integrating Bayesian networks with classical machine learning approaches lead to effective modeling of the level of mental health.

**Results:** Overall, females are more stressed than males, and people of age-group 18-29 are more vulnerable to anxiety than other age groups. Using the Bayesian Network Model, we found that people with chronic medical condition of mental illness are more prone to mental disorders during the COVID age. The new realities of working from home, home-schooling, and lack of communication with family/friends/neighbors induces mental pressure. Financial assistance from social security helps in reducing mental stress during COVID generated economic crises. Finally, using supervised ML models, we predicted the most mentally vulnerable people with ~80% accuracy.

**Question:** What factors could affect mental health, and how could this level be predicted using the same factors during the of COVID-19?

**Finding:** In this survey-data based study, multiple factors such as Social isolation, digital communication, working, and schooling from home, were identified as crucial factors of mental illness during Covid-19. Interestingly, behavioral changes such as wearing a mask and avoiding restaurants and public places were not found to be associated with mental health.

**Meaning:** Regular non-virtual communication with friends and family, healthy social life and social security are key factors and especially taking care of people with mental disease history is even more important.

## Introduction

After 7 months of initial reporting, the coronavirus pandemic continues to rage across the world. The mental health consequences of COVID-19 pandemic are profound. More than half a million lives and more than 400 million jobs have been lost^1^ causing a considerable degree of fear, worry, and concern. These effects are seen in the population at large and may be pronounced among certain groups in particular, such as youth, frontline workers, caregivers, and people with chronic medical conditions. The new world order has introduced unprecedented interventions of country-wide lockdowns that are necessary to control the spread but have led to increased social isolation. Loneliness, depression, harmful alcohol, drug use, and self-harm or suicidal behavior are also expected to rise.

The Lancet Psychiatry^2^ recently highlighted the needs of vulnerable groups during this time, including those with severe mental illness, learning difficulties, and neurodevelopmental disorders, as well as socially excluded groups such as prisoners, the homeless, and refugees. Calls to action highlighting the need for engaging more early-career psychiatrists ^3,4^, technology such as Telepsychiatry, and highlighting the high susceptibility of frontline medical workers themselves^5^ highlights the magnitude of the problem. Further, interventions are expected to have a gender-specific impact with women more likely to be exposed to additional stressors related to informal care, already existing economic disparity, and school closures. Similarly, age and comorbidity status may have a direct impact on susceptibility to mental health challenges due to their relationship with COVID-19 morbidity and mortality. Indeed, it has been established that emotional distress is ubiquitous in affected populations — a finding certain to be echoed in populations affected by the Covid-19 pandemic^6^. Finally, the role of social media is complex with some research indicating the association of social media exposure with a higher prevalence of mental health problems^7^.

However, most of these effects have been studied in isolation with a lack of modeling the collective impact of such factors. This study addresses this gap through the use of Bayesian networks, an explainable artificial intelligence approach that captures the joint multivariate distribution underlying a large survey data collected across the United States. We also address the gap of prediction of vulnerability to mental health events such as anxiety attacks using supervised machine learning models.

## Methods

### Datasets

We extracted data of 17764 adults (https://www.covid-impact.org/)^8^ from weekly surveys of the U.S. adult household population nationwide for 18 regional areas including 10 states (CA, CO, FL, LA, MN, MO, MT1, NY, OR, TX) and 8 Metropolitan Statistical Areas (Atlanta, Baltimore, Birmingham, Chicago, Cleveland, Columbus, Phoenix, Pittsburgh). To summarize, the dataset comprised variables on physical health, mental health, Insurance related Policy, economic security, and social dynamics. Table-1 shows the socio-demographic characteristics of respondents participating in the survey. Further details are available in the original source^8^.

**Table 1:**
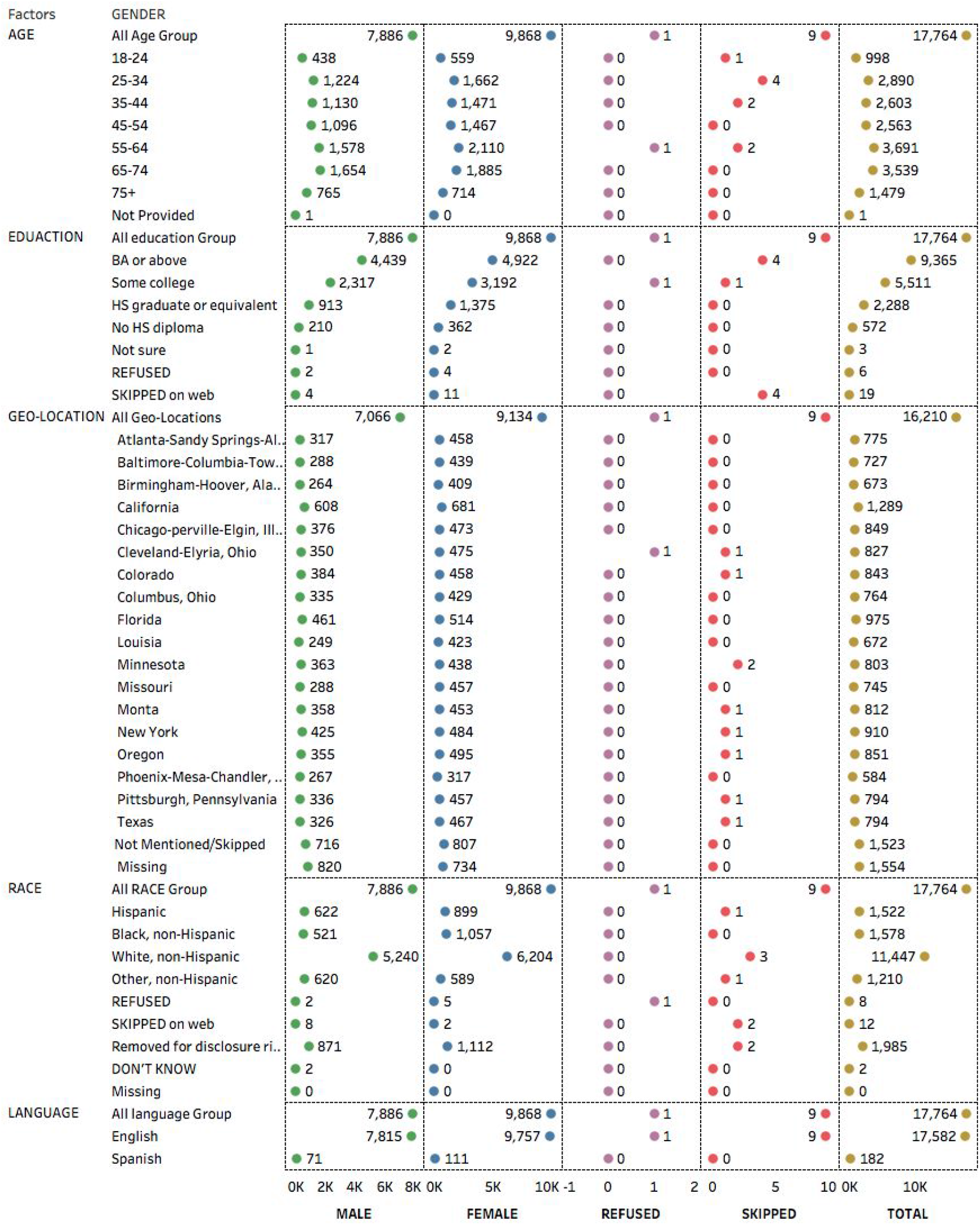
Socio-Demographics of Respondents Participated in the Survey.

### Analysis

Figure 1(a) shows the flow diagram for the analyses conducted. The survey questions were classified into several types of indicators such as mental-health, work-from-home, communication, COVID-symptoms, chronic-medical-conditions, behavioral-aspects, insurance-assistance, and many others *(see*. Table-2). We constructed a model for mental-health indicators, *soc5a* (Felt nervous, anxious, or on edge), *soc5b* (Felt depressed), *soc5c* (Felt lonely), *soc5d* (Felt hopeless about the future), and soc5e(sweating, trouble breathing, pounding heart, etc, in the last seven days) as outcome variables. Hence we first evaluated the consistency in answers to mental-health questions using Item Reliability Analysis. A scale for reliability measure of internal consistency, **Cronbach’s alpha**, was calculated using the *Psych* package in R^9^. Thereafter, a pairwise chi-square test of independence was performed to examine associations of *mental health indicators* and other variables and a p-value of less than 0.05 was taken as the cut-off for significance.

**Figure1:**
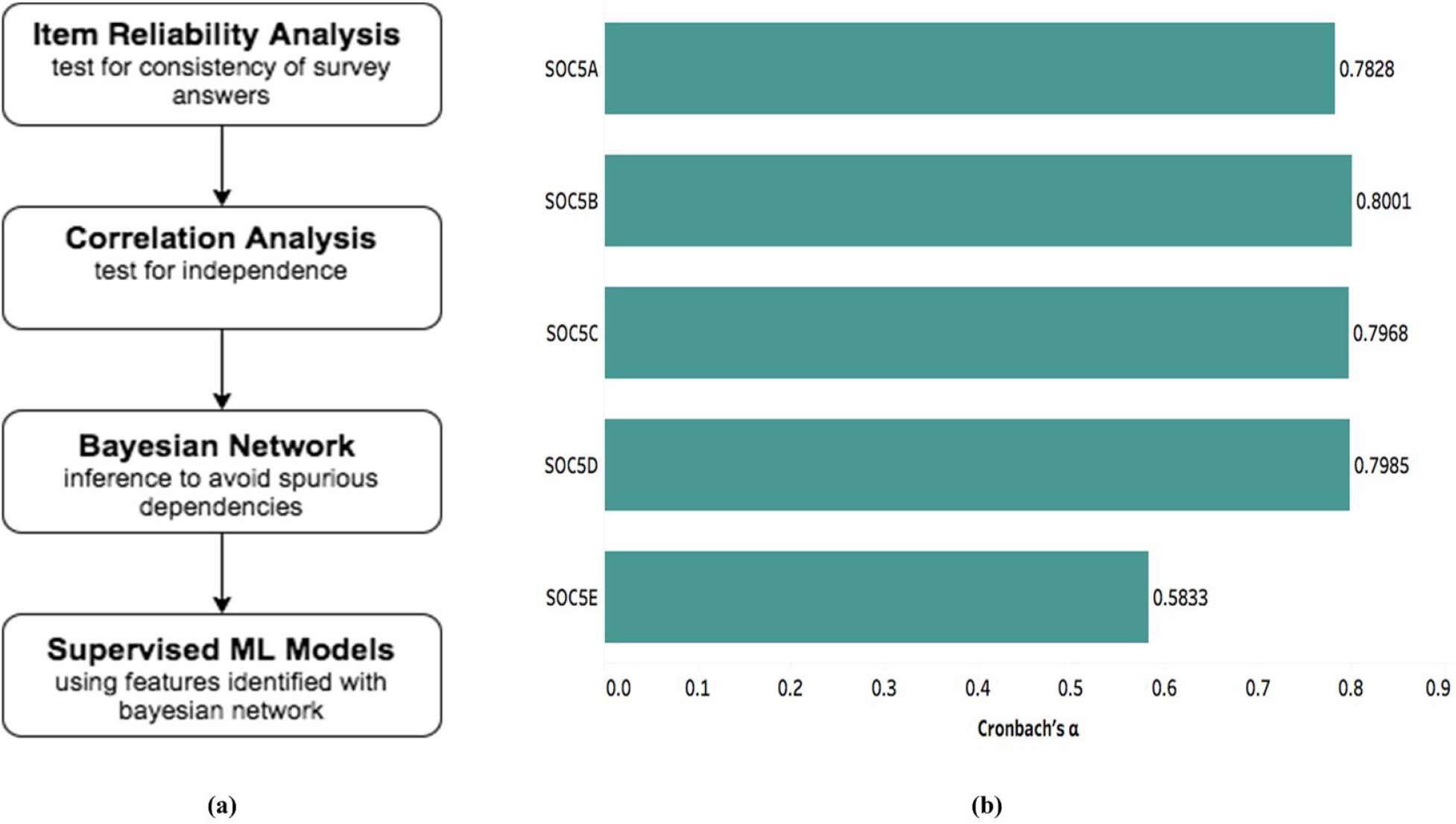
a) Outline of the analytical pipeline. b) Item Reliability Analysis of Mental Health Indicators revealed a high degree of internal consistency (Cronbach’s alpha value >0.70) for most of the psychological variables, thus indicating suitability for the modeling exercise.

**Table 2:**
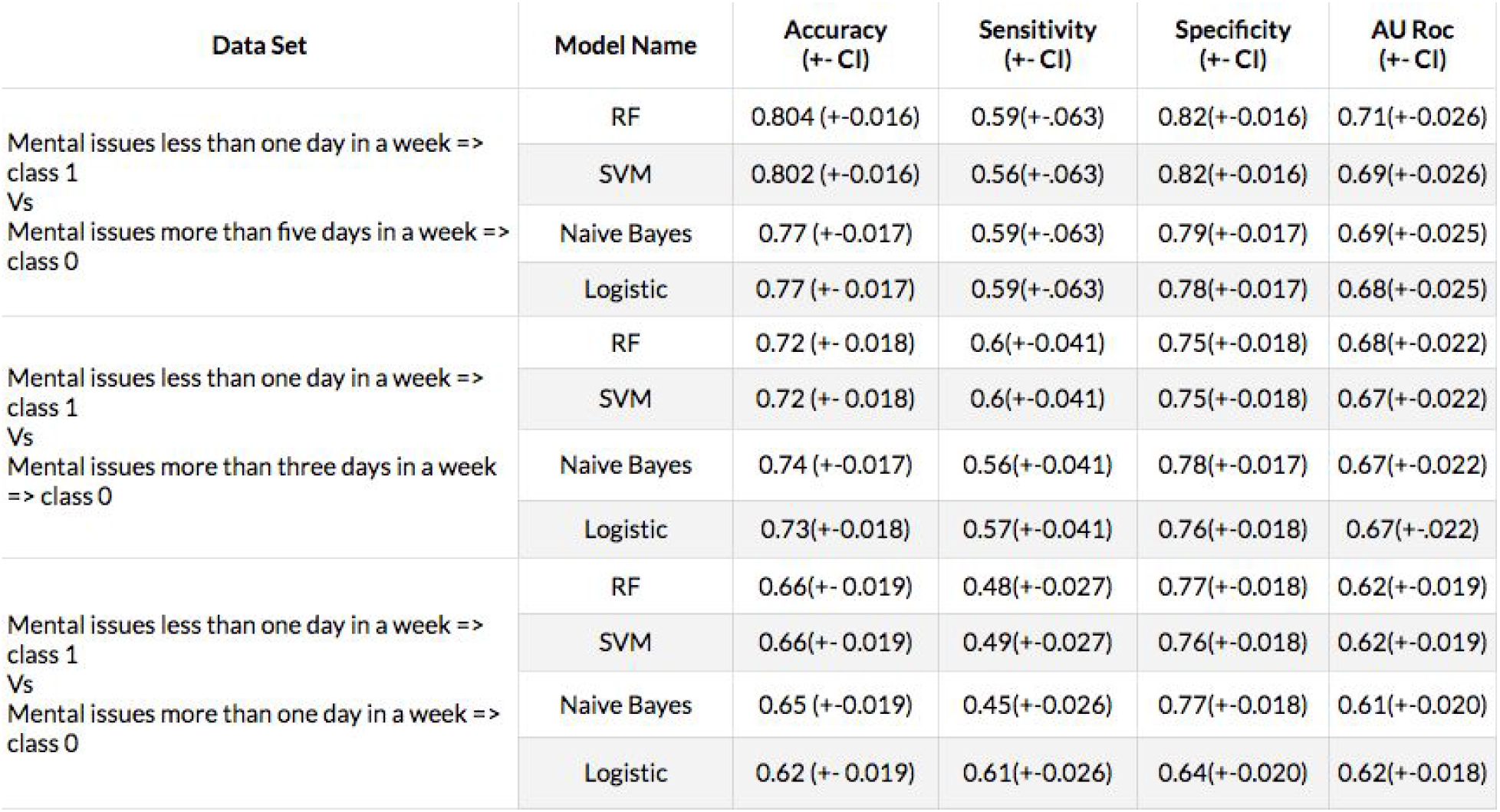
Model Performance Indicators of the Supervised Model for Prediction of Stress.

Since mental health variables may have complex dependencies with potential confounding factors, mediation, and inter-causal dependency, we extended our association analysis with data-driven Bayesian network (BN) structure learning. The structure of the learned Bayesian network was made robust through bootstrapping and ensemble averaging of edge directions. Hill climbing optimizer ^10^ with Akaike Information Criterion(AIC)^11^ based score was used to select the best PGM that explained the data. Bootstrapped learning and majority voting over 101 BNs were done. Exact Inference using the belief propagation algorithm^12^ was learned in order to quantify the strength of learned associations. The Analysis was performed in the R using the package *wiseR*^13^ Next, the *Markov-blanket*^14^ of *mental-health-indicators* was extracted to select features that may predict responses to the *mental-health-indicators*. Data were partitioned into training (80%) and testing (20%) sets and the class imbalance was corrected using the Synthetic Minority Oversampling Technique(SMOTE)^15^. Different supervised machine learning models - Random-Forest(RF), Support vector machine (SVM), logistic, naive-Bayes, were learned for predicting the response to mental health indicators using the Scikit-learn library ^16^in Python.

## Results

### Gender and age-related variation in mental-health indicators

*soc5a* (Felt nervous, anxious, or on edge), *soc5b* (Felt depressed), *soc5c* (Felt lonely), *soc5d* (Felt hopeless about the future) achieved a Cronbach’s alpha approximating 0.8 [Figure 1(b)], thus confirming their internal consistency and suitability for modeling. Gender and age-specific difference was observed in *soc5a*, with females having a higher incidence than males (two proportion z-test, p-value < 2.2e-16) (Figure 2a), and young adults in the 18-29 age group (p-value < 2.2e-16) (Figure 2b). Age group 18 - 29 in both genders was most vulnerable to mental stress > 5 days in a week, thus indicating that COVID-19 may have disproportionately affected the mental health of youth due to a variety of factors.

**Figure2:**
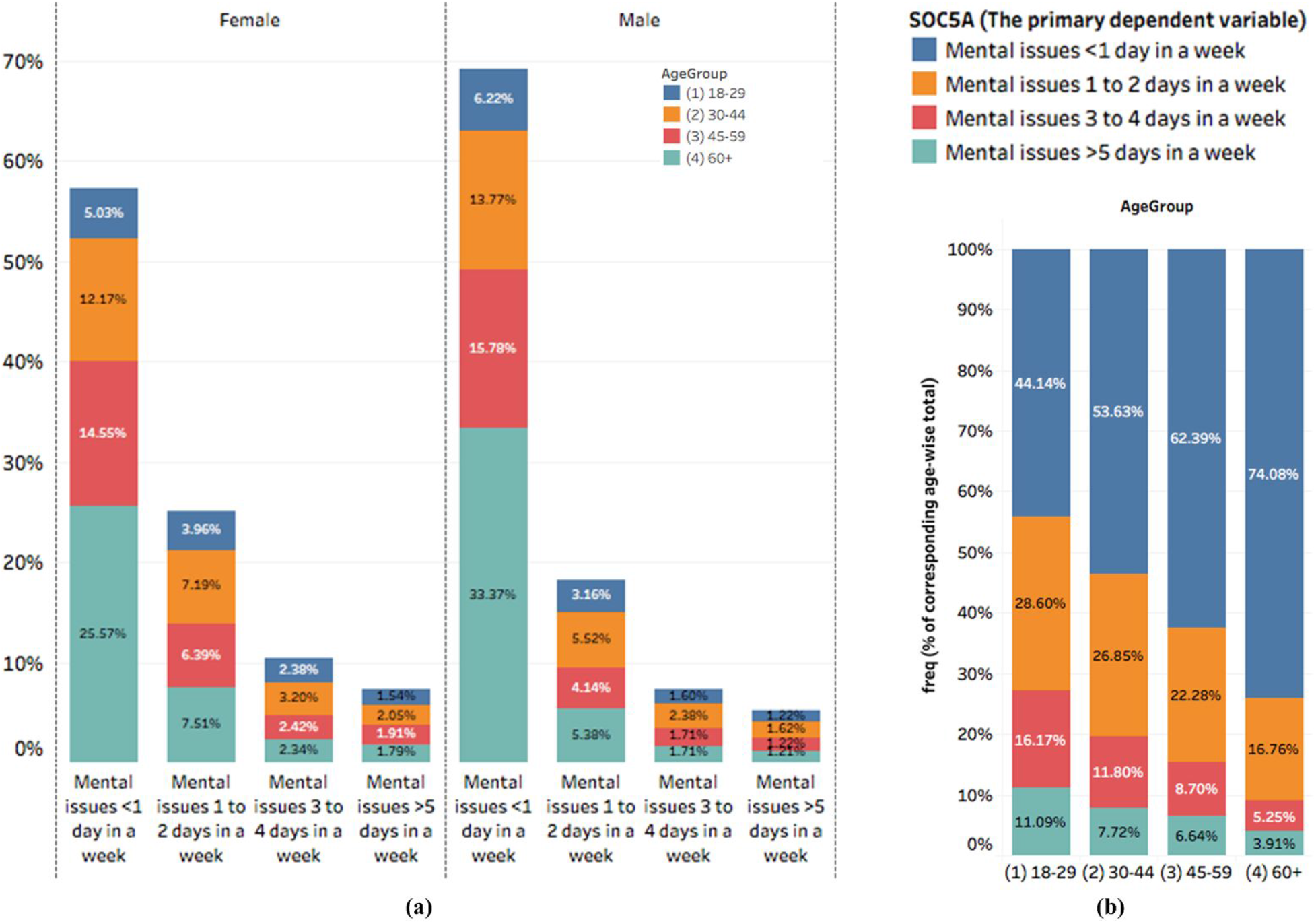
(a) Gender-wise and (b) age-wise distribution of Mental Issues (soc5a) variable. Significance was tested using two proportion z-test and chi-square test respectively, showing a higher prevalence of mental issues among youth and in women.

### Associations of anxiety in the United States

A Chi-square test revealed many significant associations of the mental health variables(Supplementary Figure 1). However, this analysis does not account for potential confounding or “explaining away” effects. Hence a data-driven Bayesian network structure-learning exercise was carried out and revealed interesting findings. From the learned structure, *soc5a* (Felt nervous, anxious, or on edge in the last seven days) was found to be the parent variable for other mental health indicators in almost 100% of the bootstrapped networks, represented as the strength of the edges [Figure 3(b)]. Being a driver variable in the structure, *soc5a* was taken as the primary dependent variable for downstream modeling analysis.

**Figure 3:**
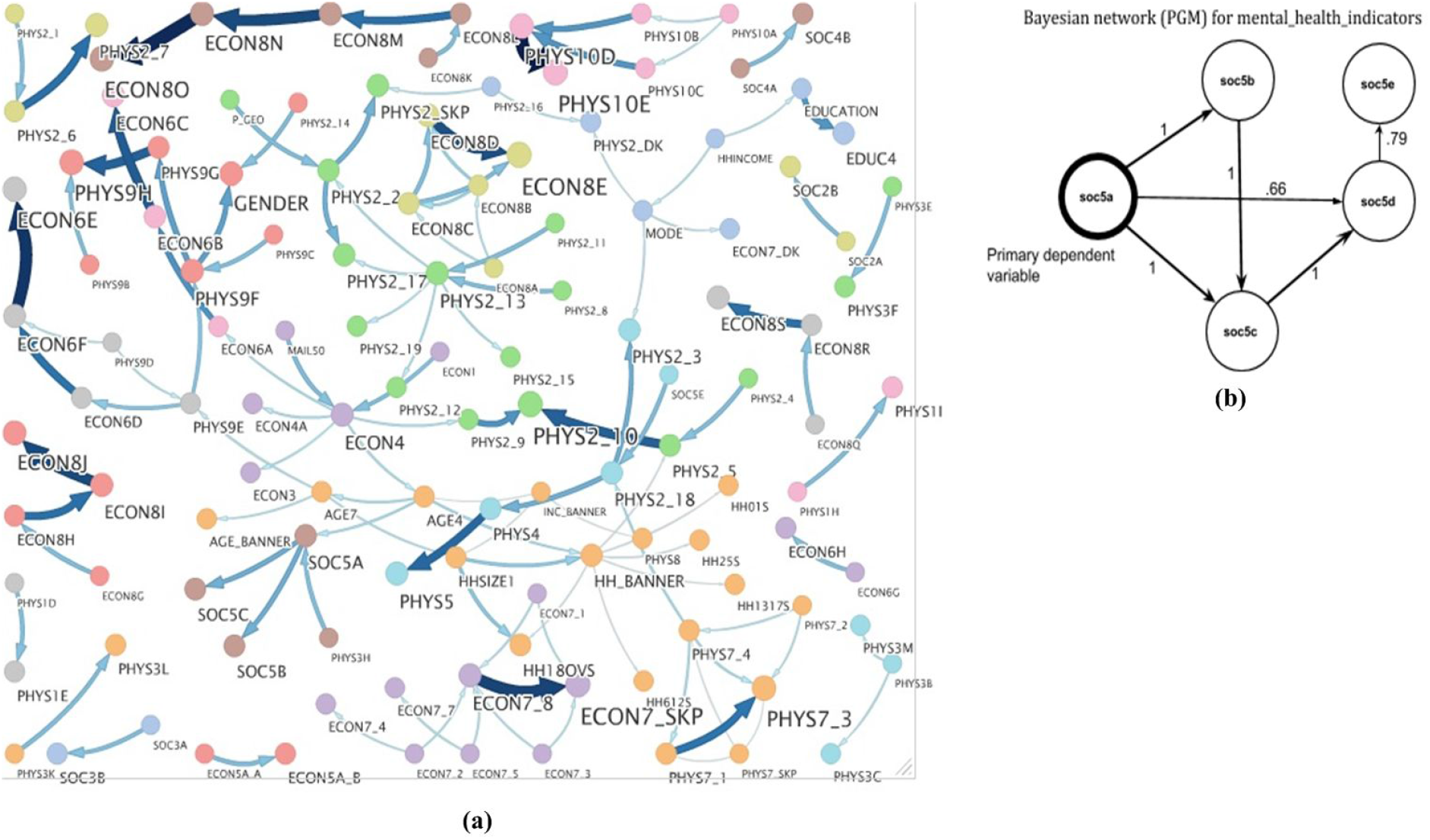
(a) Consensus structure learned through 101 bootstrapped samples. Hill-climbing search along with Bayesian Information Criterion were used to learn the structures and connections having edge strength and direction strength more than 90% are shown; (b) soc5a was found to be the parent Node of all other Mental Health Variables, therefore, leading to our choice of this variable as the Primary dependent variable.

### Impact of social life and work-related stressors

Our analysis using network inference via the Exact Inference algorithm showed a clear impact of in-person social communication on the reduction of anxiety level. A strong (>5% with CI ~1% at both sides) and (>6.5% with CI ~1.5% at both sides) monotonic increase between control of anxiety and frequency of speaking with neighbors *(soc2a, soc2b)* were observed. This effect was weaker (~1.5% with a wide confidence interval) with digital communication with friends and family conducted over phone, text, email, or other internet media (*soc3a, soc3b*). This finding underscores the importance of social communication while maintaining the appropriate measures such as masks and social distancing in order to maintain mental health during such isolating times. We also observed that the presence of kids in the house reduces the probability of depression by >11% with CI ~2% on both sides. Further, Exact Inference upon the network revealed an increase in the conditional probability of anxiety (*soc5a*) arising from canceled or postponed work (>4% with CI ~1.4%), canceled, or postponed school(7% with CI ~1.5%), worked from home(>5% with CI ~1.3) and studied from home(>7% with CI ~1.8). Interestingly, although 83% of all volunteers chose to wear the mask, 77% avoided restaurants, 83% avoided public and crowded places these measures were not found to be associated with a significant change in anxiety levels as inferred from our model. These inferences are summarized in figure-4.

**Figure-4:**
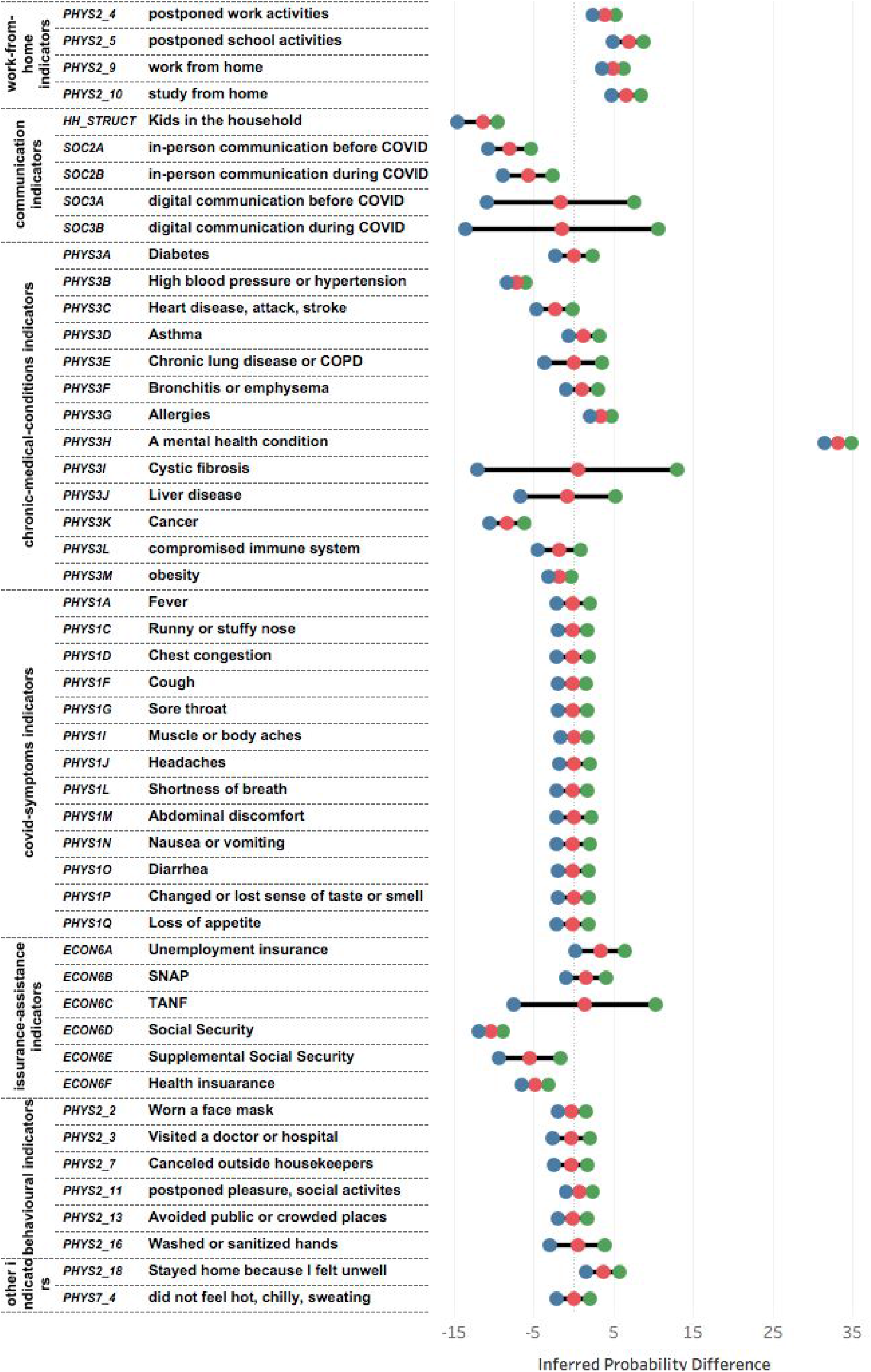
Inferences from the Bayesian network: The difference in inferred probability was calculated after conditioning on the independent variables. A positive association implies a mental-stress-inducer whereas a negative association implies mental-stress-reducer. The red circle shows the mean value with green and blue showing confidence intervals.

### Impact of symptoms and comorbidities

We also investigated the relationship between mental stress and COVID symptoms indicators. WHO recommends contacting health service providers if any COVID symptoms (*phys1a* to *phys1q*) are experienced within the last seven days. Our network did not indicate any significant impact of these responses on mental health (*soc5a*), the conditional probability of which remained unchanged (62.2%) across the responses. Surprisingly, although medical conditions *(phys3a* to *phys3m)* are known to increase the risk of serious illness from COVID-19, our model showed that suffering from cancer *(phys3k*) and hypertension *(phys3b*) had a reverse impact on anxiety levels. Those suffering from cancer had approximately 8.3% (with ~2% CI) higher conditional probability of having less than one anxiety-ridden day in a week (>7% effect for hypertension with CI ~1.5%). Additionally, Cystic-fibrosis (phys3i) and Liver-disease (phys3j) have wide confidence intervals with non-significant differences in mean values. (Figure-4).

### Impact of economic factors

Receiving income assistance through Social Security improved the conditional probability of less than one day of anxiety in a week by 10.4% (with CI ~1.5%) as compared with the segment of people who did not apply or receive it. Just applying for income assistance led to a 4% improvement (Figure-4). Similarly, Supplemental Social Security (~5.5 with CI ~4%) and Health insurance (~5% with CI ~2%) also led to similar results.

In addition to this, Old age people found health insurance more relaxing than young age people. COVID has also severely affected the financial condition of individuals. That may also lead to mental stress.

### Predictive modeling for susceptibility to anxiety attacks

Our supervised modeling approach used the Markov blanket of the *soc5a* variable, i.e. age (*age4)*, physical symptoms in the last seven days (*phys7_4*), stay at home (*phys2_18*), and prior clinical diagnosis of any mental health condition (*phys3h*) as predictors.

The following three prediction scenarios were considered-

1. Mental issues *less than one day* in a week (class 1) vs. Mental issues *more than* ***one*** *day* in a week (class 0)
2. Mental issues *less than one day* in a week (class 1) vs. Mental issues *more than* ***three*** *days* in a week (class 0)
3. Mental issues *less than one day* in a week (class 1) vs. Mental issues *more than* ***five*** *days* in a week (class 0)

We observed a decay (Accuracy 0.80 to 0.64, also confidence intervals mentioned in the table-2) in model predictability as we moved from high risk of depression (case-3) to low risk of depression (case-1) [figure4]. Such a trend was visible with all 4 machine learning techniques we used. Our ML properties (accuracy, sensitivity, specificity, AUROC) are summarized in table-2.

## Discussion

Mental Health is a serious public health concern. Mood disorders and suicide-related outcomes have increased significantly over the last decade among all age groups and genders^17 18^. The rapid spread of coronavirus infection forced governments across the world to close public gathering places, schools, colleges, restaurants, and industries. Social isolation, digital communication, working, and schooling from home have become the new normal and many jobs have been lost. Collectively, this has triggered a high level of anxiety, stress, and depression, globally. We did not find studies that have used models to not just predict but also to explain the subtle effects of life-situations on mental health. An explainable probabilistic graphical modeling approach with bootstraps and exact inference allowed us to capture many of these effects in a robust manner. Our study revealed that individuals having a prior diagnosis of any mental illness are the most vulnerable for mental illness during the COVID-19 phase, which recommends building national-level policies to regularly track their mental status and treat them accordingly. Most importantly, our results re-iterate the economic underpinnings of collective mental health response. Income assistance via Social Security or Supplemental Social Security had a demonstrable effect on the alleviation of anxiety as inferred from our model, which provides the first scientific evidence, to the best of our knowledge, proving the utility of such efforts. The effect of the extent of such measures may be captured in such modeling studies conducted in various parts of the globe, with widely varying assistance structures during this time.

Our findings from the United States can also stimulate further cultural and social research in other geographies with similar or different social structures. For example, the effects of in-person communication, as opposed to digital connectedness, may be different in countries where community living and joint families are still commonplace, e.g. India. Digital connectedness was not as effective as talking to a neighbor, at least in the United States highlighting that these are fundamentally different influences on mental health and need to be further explored in systematic studies. We conjecture that such differences may arise from the evolutionary mechanisms that have shaped human societies to live and share in close physical connectedness. Such an effect has been previously shown in primates kept in isolation who display depressive symptoms ^19 20^. Similarly, parenting and its association with neuropeptide hormones may partially explain ^21^ our results that the presence of kids reduces anxiety levels. Interestingly, the COVID-19 pandemic has created a unique natural experiment on the collective mental health response of individuals to a health emergency.

The life-cycle of such a response may need to be further studied as the world goes through various phases of the pandemic to its resolution. However, our study indicates that the mental health impact is observable within a span of a few months, especially on young individuals. Further research will be needed, ideally in a longitudinal setting, where the same individuals can be surveyed again to understand the dynamics of the collective mental health response.

Our results also highlight that modern technological development in virtual communication is not able to replace natural socializing. Hence it becomes imperative to design better and empathetic technological tools that may shape a society that prevents isolation and alienation even while maintaining physical distancing and preventive measures for limiting spread. Personalization and contextualization of such measures will also be important as our results indicate that persons with previous mental health conditions may be disproportionately affected. Finally, our results indicate that it may be possible to identify people at the highest risk of developing mental health disturbances. Our model achieved its best performance for those who were most vulnerable (having mental stress more than five days in a week) vs. least vulnerable people (having no stress or less than one-day stress in a week). This can help in the segmentation of vulnerable populations such as front-line healthcare workers and who are facing disproportionately higher levels of stress during this time.

We could not explain why anxiety levels may be lower in persons with pre-existing cancer or hypertension. This may be a result of reduced work-environment related stress or more contact with family members at home. However, the current dataset is not suited to address this at a finer level of explainability. Our study has a few limitations. The dataset used for training the model is cross-sectional and we could not comment upon the temporality and persistence of the discovered effects. Secondly, our results were currently limited to only one geography, i.e. the United States. However, the relatively large sample size and multi-ethnic involvement in the survey allowed us to construct a robust network with ensemble averaging and majority voting from 101 bootstrapped networks. Therefore, the discovered influences are likely to hold true in the United States. Further, we believe that this is the first attempt to quantify the impact of social factors on mental health through an explainable AI model and many of our findings are intuitive. The ranking of features and quantification of this impact couldn’t have been intuitively achieved without modeling and this study will provide a basis for many further studies and design of effective social interventions to mitigate the mental health impact of natural health disasters.

## Data Availability

NA

https://www.covid-impact.org/

## Author Contribution

Study Design: VK,TP,IJ, Dataset: IJ,AK, Data Analysis: IJ,RA, Paper writing: IJ,RA,AK,TP Paper Review: VK,TP

---

**Supplementary Table 1:**
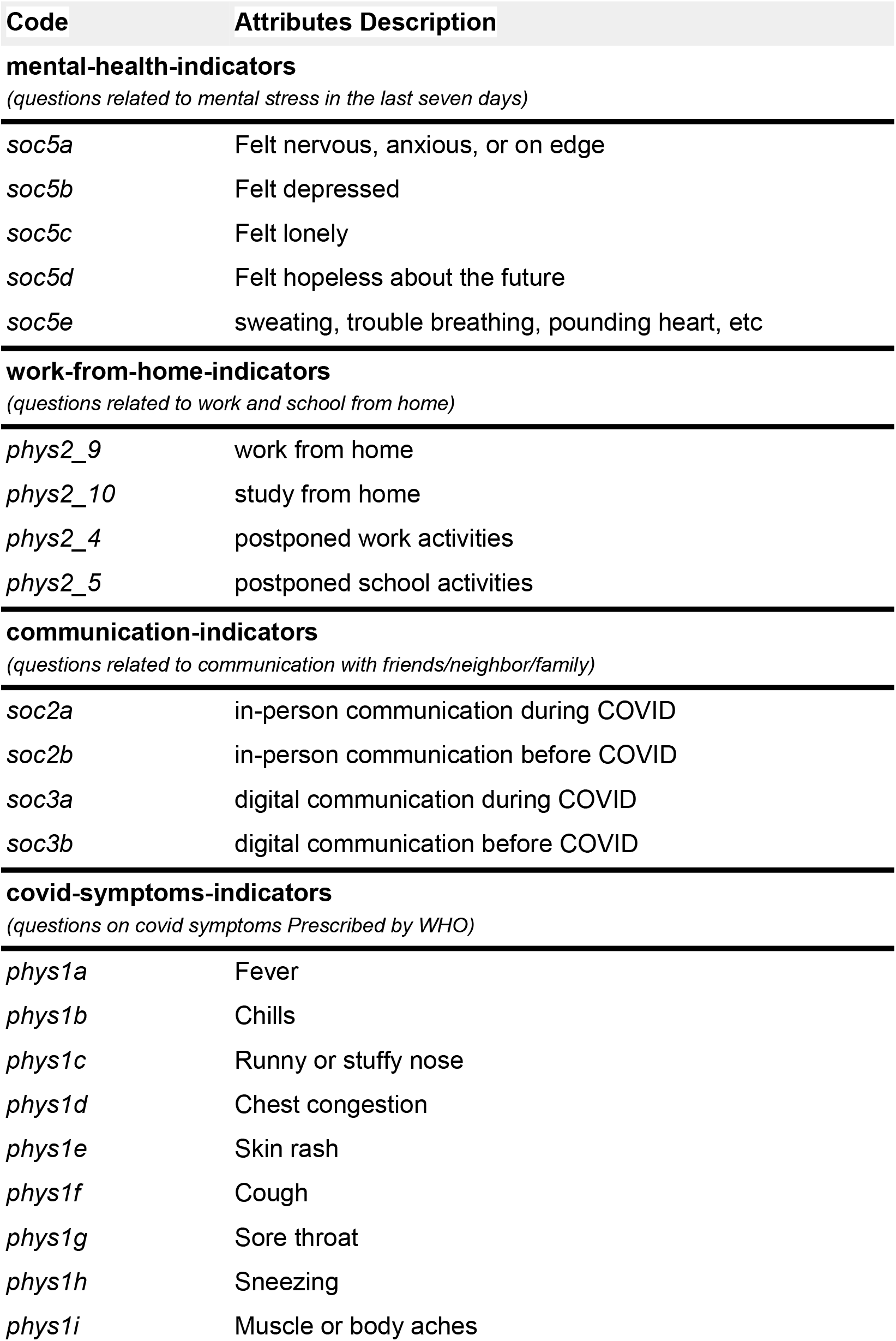
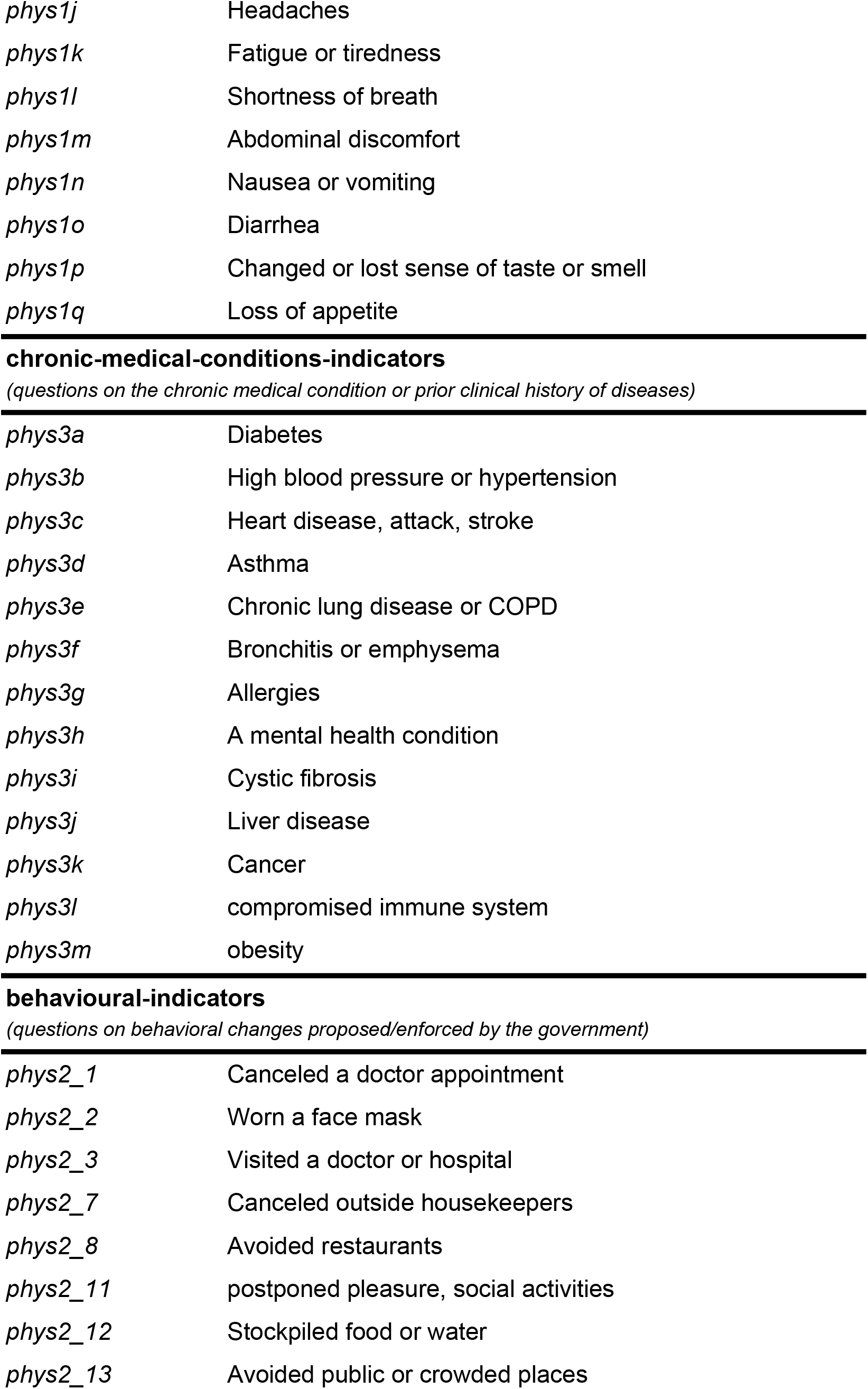
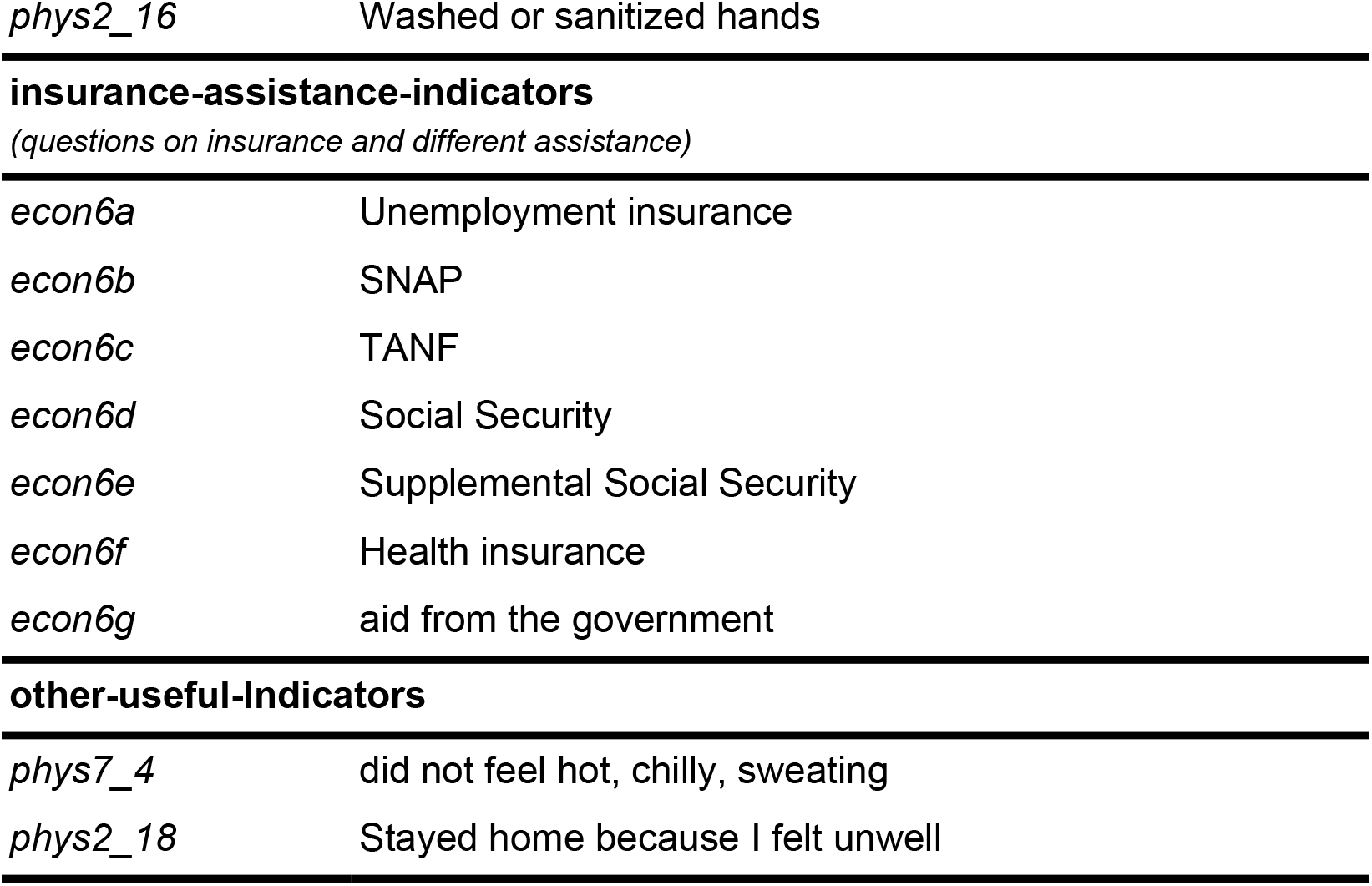
variable groups as Indicators

| Code | Attributes Description |
| --- | --- |
| **mental-health-indicators** |  |
| *(questions related to mental stress in the last seven days)* |  |
| *soc5a* | Felt nervous, anxious, or on edge |
| *soc5b* | Felt depressed |
| *soc5c* | Felt lonely |
| *soc5d* | Felt hopeless about the future |
| *soc5e* | sweating, trouble breathing, pounding heart, etc |
| **work-from-home-indicators** |  |
| *(questions related to work and school from home)* |  |
| *phys2_9* | work from home |
| *phys2_10* | study from home |
| *phys2_4* | postponed work activities |
| *phys2_5* | postponed school activities |
| **communication-indicators** |  |
| *(questions related to communication with friends/neighbor/family)* |  |
| *soc2a* | in-person communication during COVID |
| *soc2b* | in-person communication before COVID |
| *soc3a* | digital communication during COVID |
| *soc3b* | digital communication before COVID |
| **covid-symptoms-indicators** |  |
| *(questions on covid symptoms Prescribed by WHO)* |  |
| *phys1a* | Fever |
| *phys1b* | Chills |
| *phys1c* | Runny or stuffy nose |
| *phys1d* | Chest congestion |
| *phys1e* | Skin rash |
| *phys1f* | Cough |
| *phys1g* | Sore throat |
| *phys1h* | Sneezing |
| *phys1i* | Muscle or body aches |
| *phys1j* | Headaches |
| *phys1k* | Fatigue or tiredness |
| *phys1l* | Shortness of breath |
| *phys1m* | Abdominal discomfort |
| *phys1n* | Nausea or vomiting |
| *phys1o* | Diarrhea |
| *phys1p* | Changed or lost sense of taste or smell |
| *phys1q* | Loss of appetite |
| *phys3a* | Diabetes |
| *phys3b* | High blood pressure or hypertension |
| *phys3c* | Heart disease, attack, stroke |
| *phys3d* | Asthma |
| *phys3e* | Chronic lung disease or COPD |
| *phys3f* | Bronchitis or emphysema |
| *phys3g* | Allergies |
| *phys3h* | A mental health condition |
| *phys3i* | Cystic fibrosis |
| *phys3j* | Liver disease |
| *phys3k* | Cancer |
| *phys3l* | compromised immune system |
| *phys3m* | obesity |
| *phys2_1* | Canceled a doctor appointment |
| *phys2_2* | Worn a face mask |
| *phys2_3* | Visited a doctor or hospital |
| *phys2_7* | Canceled outside housekeepers |
| *phys2_8* | Avoided restaurants |
| *phys2_11* | postponed pleasure, social activities |
| *phys2_12* | Stockpiled food or water |
| *phys2_13* | Avoided public or crowded places |
| *econ6a* | Unemployment insurance |
| *econ6b* | SNAP |
| *econ6c* | TANF |
| *econ6d* | Social Security |
| *econ6e* | Supplemental Social Security |
| *econ6f* | Health insurance |
| *econ6g* | aid from the government |
| **other-useful-Indicators** |  |
| *phys7_4* | did not feel hot, chilly, sweating |
| *phys2_18* | Stayed home because I felt unwell |

**Supplementary Figure 1:**
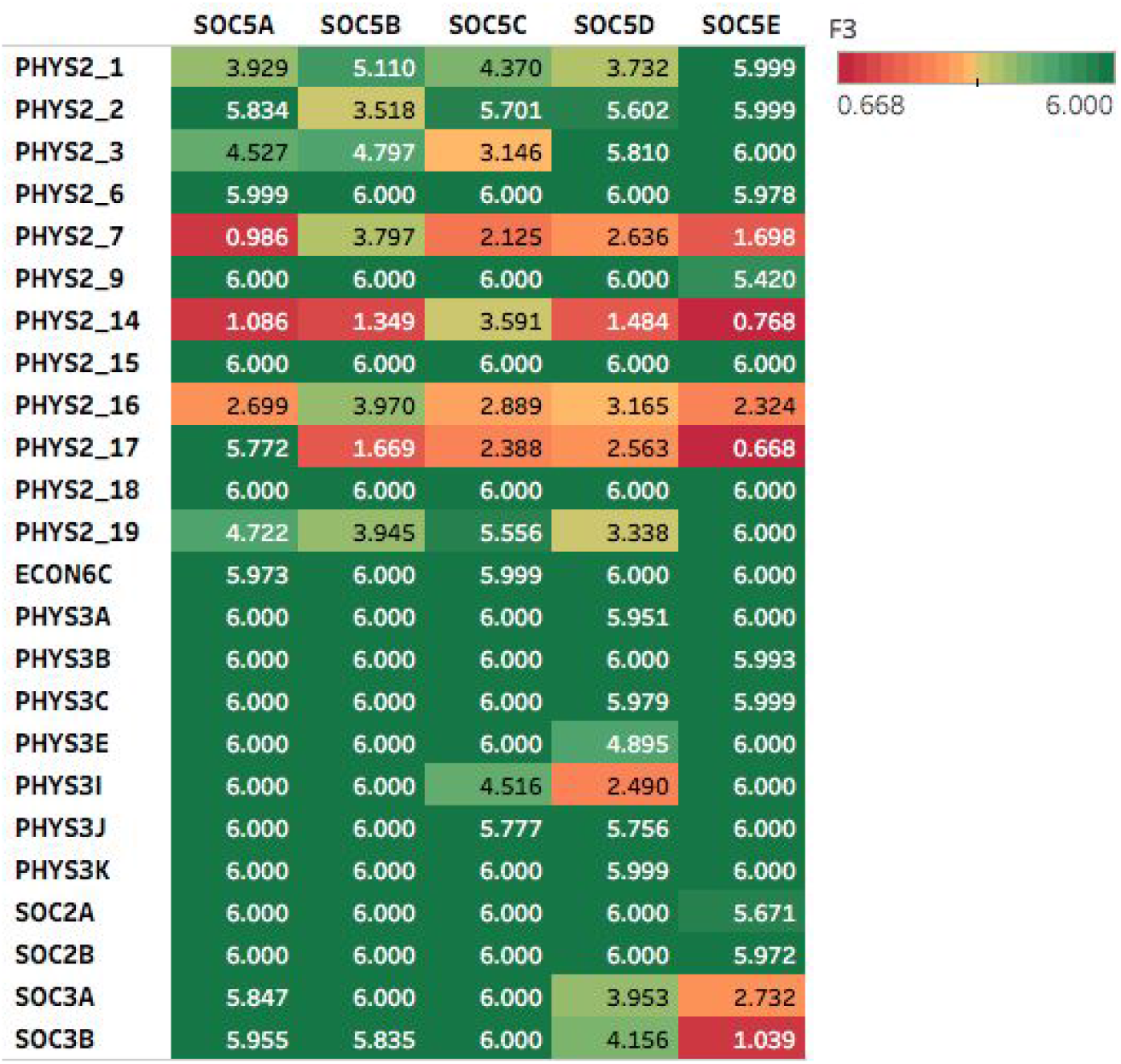
Heatmap for negative log p values for the significant association.

**Supplementary Figure 2:**
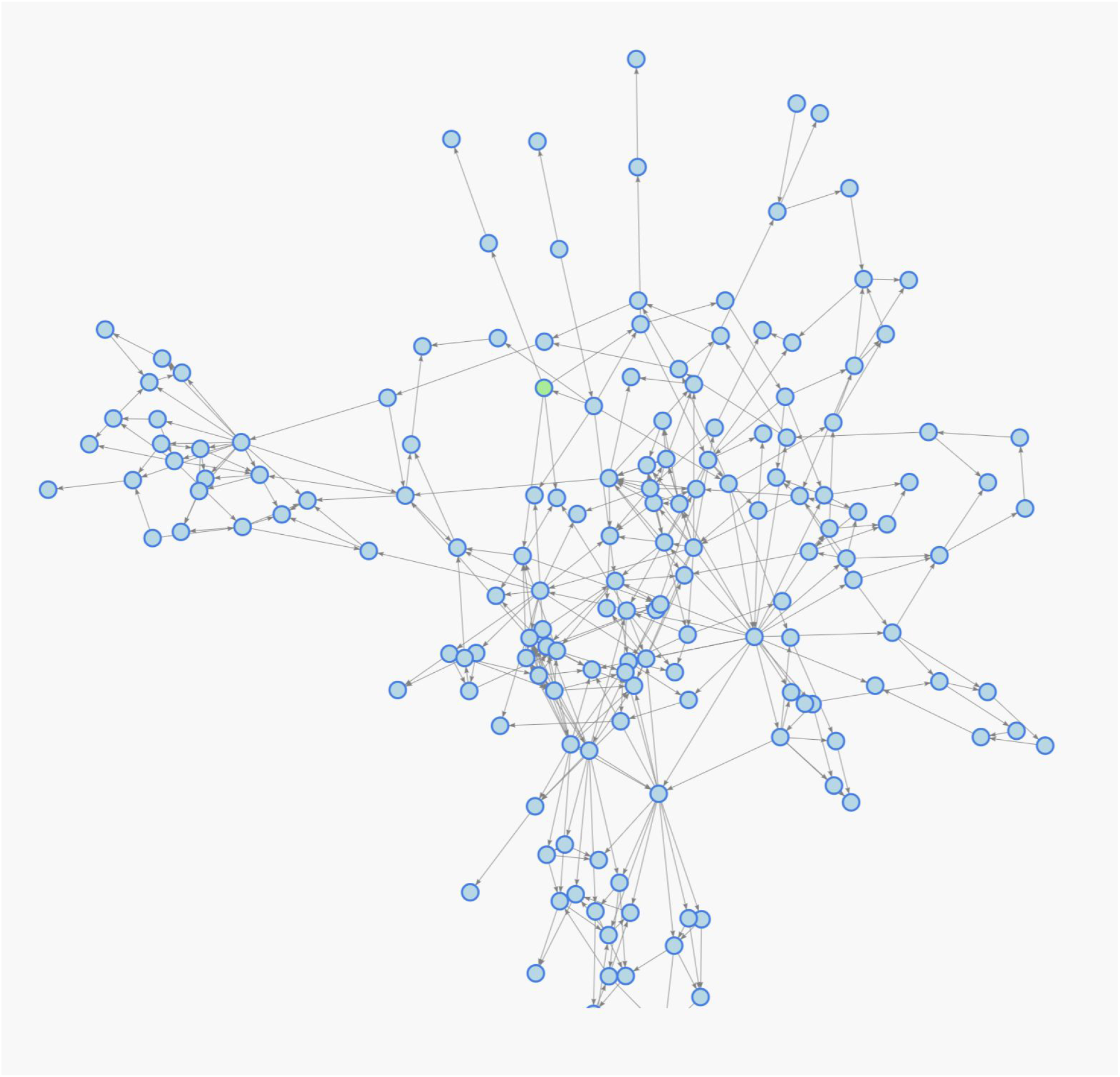
Bayesian Network topology

